# Adherence to Pakistan dietary guidelines – Findings from major cities of Pakistan

**DOI:** 10.1101/2020.07.06.20147017

**Authors:** Ibrar Rafique, Arif Nadeem Saqib Muhammad, Nighat Murad, Muhammad Kashif Munir, Aftab Khan, Rabia Irshad, Tayyaba Rahat, Saima Naz

**Author notes:** **Corresponding author:** Dr. Muhammad Arif Nadeem Saqib, Senior Research Officer, Pakistan Health Research Council, Shahrah-e-Jamhuriat, Constitution Avenue, G-5/2, Islamabad. **Authorship:** IR conceived and design the study, write proposal for grant and draft manuscript. MANS revise the proposal, interpret data, facilitate data collection, add intellectual content to manuscript. NM revised the manuscript critically for intellectual content. KM, AK and RI played a role in acquisition of data from field work in different cities. TR and SN performed data analysis and interpret the results. **Ethical Standards Disclosure:** The study was conducted as per Helsinki declaration. The Ethical Clearance was taken from National Bioethics Committee (NBC 246). Written informed consent was taken from all participants.

## Abstract

**Background:** Pakistan dietary guidelines for better nutrition were developed to cater the local need and prevent nutritional deficiency by providing information to public about healthy eating practices.

**Aims:** To assess the level of adherence to Pakistan Dietary Guidelines for Better Nutrition (PDGN)

**Methods:** It was a community based study conducted in five cities with two stage stratified sampling approach. Total of 448 participants were interviewed using Food frequency questionnaire adapted to local context. Five food groups (proteins, cereals, dairy, vegetables and fruits) were taken as per country guidelines. A score point of 1 was given to each food group making a total of 5 scores. Data were analyzed using SPSS.

**Results:** Overall adherence to PDGN was poor as none of the participants had 05 score while only 1% achieved score 04. However, adherence was more in females (B = 0.45, 95%CI = 0.24; 0.66), graduates (B = 0.45, 95% CI = 0.25; 0.64), unmarried (B = 0.30, 95% CI = 0.18; 0.43), unemployed (B = 0.22, 95% CI = 0.01-0.43) and aged >50 years (B = 0.34, 95% CI = 0.08; 0.60) as compared to others. Among food groups, mean intake of cereals (carbohydrates) was high (3.38±1.39) followed by other items with fruits was least (0.76±0.91). Overall, at least one serving of discretionary food was taken by participants which was more female gender (p= 0.001), graduates (p= 0.003), high socio-economic group (p=0.001) and employed persons (p= 0.04).

**Conclusion:** The adherence to PDGN was poor and there is a need to bring behavior change by information education and communication to the society.

## Introduction

Dietary intake is important to one’s health as an unhealthy dietleads to non-communicable diseases (NCDs) which are a leading cause of mortality in the world. It is reported that an unhealthy diet is generally associated with low education, low profession and low socioeconomic condition (1, 2).The National Dietary guidelinesare developed by the Country to improve the nutritional well being of the population by adopting and promoting healthy lifestyles and eating practices(3). The objective of the guidelines is to make recommendations about the components of a healthy and nutritionally adequate diet to help promote health and prevent chronic disease for current and future generations(4).

Pakistan is a developing country, facing a double burden of Communicable Diseases and Non-communicable diseases(5). The NCDs risk factor survey revealed that 96.5% were consuming an unhealthy diet (6). Similarly, it was reported that healthy dietary items i.e. fruits, vegetables, whole grains, nuts and minerals are being replaced with foods rich in saturated fats, refined sugar and salt(7).The country-specific guidelines (Pakistan Dietary guidelines for better nutrition PDGN) were developed by Ministry of Planning, development and reforms & Food and Agriculture Organization of United Nations to meet the nutritional requirement of the population by taking into account local cooking methods, dietary practices and health situation of the country. The PDGNis very comprehensive and providing information to the general public about healthy eating practices which ultimately will prevent them from chronic diseases(8).

## Objectives

The current study was planned to assess the dietary pattern and adherence to the national dietary guidelines (PDGN).

## Methods

### Study design and settings

It was a community-based cross-sectional survey which was conducted in 05 cities including Federal Capital (Islamabad) and Provincial Capitals (Lahore, Karachi, Peshawar and Quetta). The study was conducted in 2018-2019. From each city, three Union Councils were randomlyselected belonging to low, middle and high socioeconomic groups. The low socioeconomic area was categorized as rural area predominantly with the families having small earning. The middle socioeconomic area was defined where most of the persons owned a house &, were having a motorcycle or an economy car while high socioeconomic group having high income (>100,00 rupees) and 1300 cc car.

### Study population and sample

Adult’s ≥18 years of both genders from the study area who agreed to participate were included in the study. Mentally disabled and those living temporally in the household were excluded from the study. The sample size was calculated using the WHO sample size calculator with a 95% confidence level anda design effect of 1. Keeping in view non-response rates a total of 90 households were selected from each city making a total of 450 households. Equal number of participants were taken from low, middle and high socioeconomic groups.

### Sampling method/technique

Two-stage stratified sampling approach was used for selection of Union Council (UCs) and households i.e. selection of Union Councils and then selection of households. Within the UC, a cluster of 150 households was selected randomly from a landmark place like school, hospital, masjid or market and enumerated. Among them, 30 households were selected by taking every 5th household using systematic random sampling. The head of the house or elder person living in the household was interviewed. In case, where no one waspresent in the sampled house or the house was found locked, the team then went to the house next to the selected house.

### Data collection

The international standardized Food frequency questionnaire was adapted to the local context. It was translated into Urdu as it is the most widely spoken language in the country. The data collection team comprised of the enumerator, data collectors (one male and one female) and trained on study objective and methods. The enumerator marked the houses while the data collector interviewed the household members.

The information related to demography i.e. gender, occupation, marital status, education was taken. The food listed in Food frequency questionnaire were categorized into i) Meats ii) Milk and derivatives iii) Grains (Cereals), iv) Fruits v) Vegetables vi) Discretionary food items and other questions. The participants were asked to recall the food intake pattern and below mentioned options were given options as Daily (once a day, 2-3 per day, 4-5 per day), Weekly (once a week, 2-4 per week, 5-6 per week), Monthly (never or less than once/month, 1-3 per month).

The dietary recommendations were taken from PDGN prepared by the Planning commission of Pakistan(8). According to the PDGN, there must be daily 2-3 servings each of meat and pulses, vegetables, milk and milk products, fruits and 4-5 times cereals & grains. The adherence was calculated by taking a score of 01 for each category if the participants were taking 2 or 3 servings of each meat and pulses, vegetables, milk and milk products, fruits daily while 0 scores weregiven in case of less than 02. Similarly score of 01 was given if participants were taking cereals/grains 4 or more than 4 times and 0 scores were given in case of taking less than 04 times. A maximum score of 05 was given.

### Data Entry, management and analysis

The forms were collected and data was entered in SPSS sheet by the team of data entry operators. The entered data was compared for errors and corrected accordingly by validating the values against those in the questionnaires. The data were analyzed by using SPSS 21. The daily and weekly servings were calculated for each dietary item. The p-value less than 0.05 were considered as significant. The comparison between adherence score and different socio-demographic determinants was done by using t-test and ANOVA. The social determinants were further analyzed by using multiple linear regression method.

### Ethics Statement

The study was approved by National Bioethics Committee of the Country (NBC 246). Written informed consent was taken from all the participants prior to enrolment.

## Results

Of the total, 54% were female and 46% male with 41% were undergraduates followed by 29% graduate and above. Almost 41% had passed primary or less than primary. Almost 47% had either government or private job and 15% had their own business while were 26% housewives and 12% were unemployed.

Overall adherence to dietary guidelines (PDGN) was poor as none of participants achieved maximum score i.e. 05, only 1% achieve the score of 04, 10% had score 03, 28% had 02 score, 36% had score 1 while 25% had none score. The analysis showed that adherence was significantly high among females (B = 0.45, 95% CI = 0.24; 0.66), graduates (B = 0.45, 95% CI = 0.25; 0.64), unmarried (B = 0.30, 95% CI = 0.18; 0.43), unemployment (B = 0.22, 95% CI = 0.01-0.43) and those aged age >50 years (B = 0.34, 95% CI = 0.08; 0.60) (Table 1).

**Table 1.**
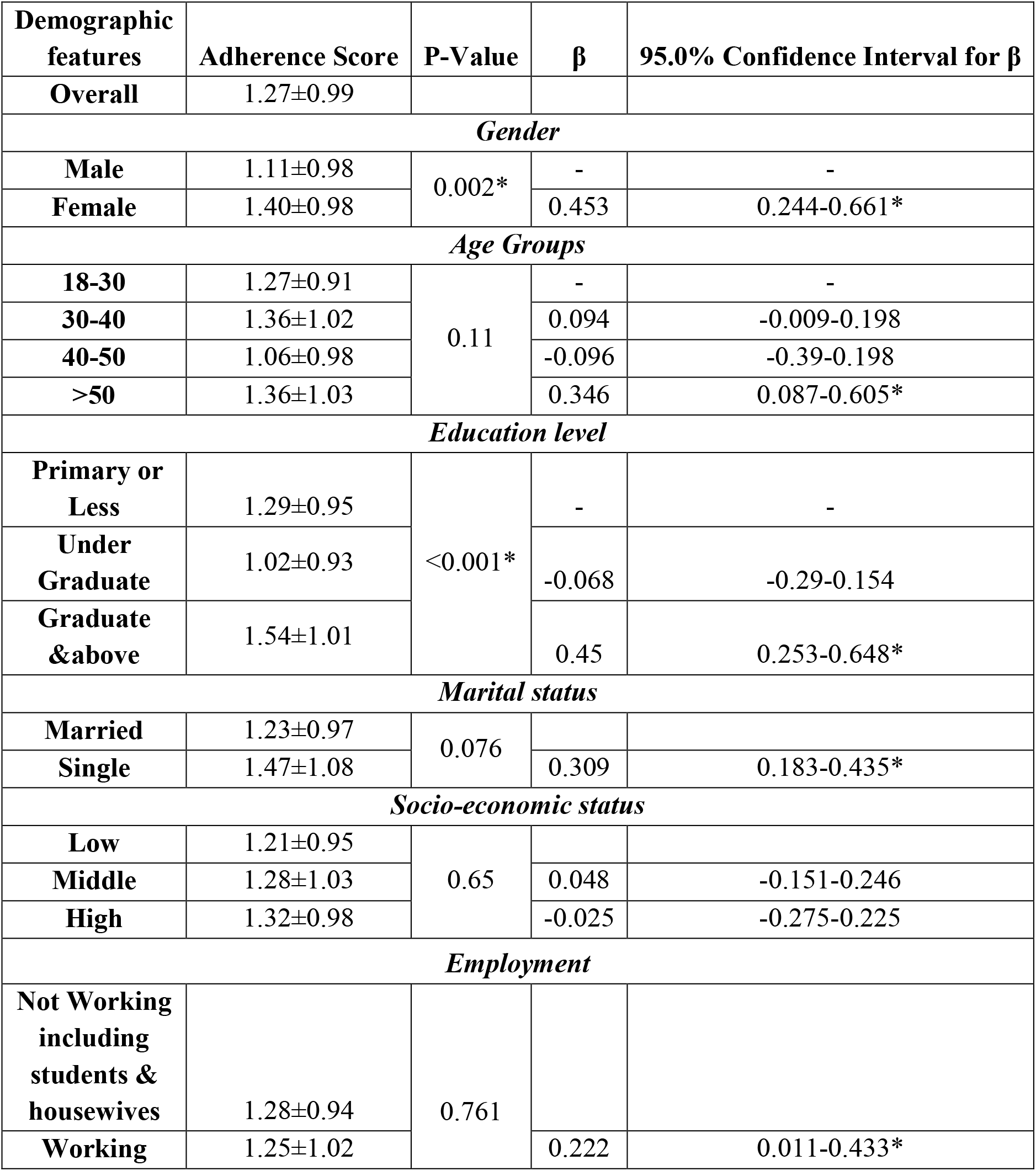
Association of adherence score of Pakistani adults with socio-demographic determinants:

Among the five food groups, the daily intake or serving of carbohydrates was high i.e. 3.38 followed by Milk & derivatives (2.46), vegetables (1.47), proteins (1.18) and fruits (0.76). No association was seen between the daily intake of carbohydrates, proteins and vegetables and demographic characteristics. However, the intake or daily serving of dairy was significantly high among female (p=0.001) and in the age group 30-40 years (p=0.02) while the daily fruit intake/serving was significantly common in high socio-economic groups (p=0.028) (Table 2). The use of discretionary food items was higher among female gender (p= 0.001), graduates (p= 0.003), high socio-economic group (p=0.001) and employed persons (p= 0.04). (Table 3)

**Table 2.** Daily mean intake of food groups with socio-demographic features:

| **Food Groups** | **Carbohydrates** | **Proteins** | **Dairy** | **Vegetables** | **Fruits** |
| --- | --- | --- | --- | --- | --- |
| **Overall** | 3.38±1.39 | 1.18±0.71 | 2.46±1.75 | 1.47±2.24 | 0.76±0.91 |
| ***Gender*** |  |  |  |  |  |
| **Male** | 3.19±1.18 | 1.12±0.49 | 1.98±1.47 | 1.31±2.39 | 0.69±0.88 |
| **Female** | 3.54±1.54 | 1.23±0.84 | 2.87±1.86 | 1.6±2.09 | 0.82±0.92 |
| **P-Value** | **0.007** | **0.11** | **<0.001** | **0.119** | **0.162** |
| ***Age Groups*** |  |  |  |  |  |
| **18-30** | 3.39±1.17 | 1.3±0.863 | 2.63±1.74 | 1.46±2.34 | 0.73±0.83 |
| **30-40** | 3.37±1.45 | 1.17±0.55 | 2.75±1.96 | 1.54±2.14 | 0.84±1.00 |
| **40-50** | 3.25±1.31 | 1.09±0.51 | 2.22±1.63 | 1.19±2.03 | 0.72±0.77 |
| **>50** | 3.48±1.52 | 1.09±0.40 | 2.17±1.33 | 1.61±2.72 | 0.64±0.71 |
| **P-Value** | **0.743** | **0.089** | **0.028** | **0.615** | **0.421** |
| ***Education Level*** |  |  |  |  |  |
| **Primary** | 3.54±1.23 | 1.21±0.63 | 2.64±1.84 | 1.60±2.58 | 0.70±0.70 |
| **Undergraduate** | 3.24±1.29 | 1.14±0.58 | 2.32±1.79 | 1.37±2.46 | 0.67±0.73 |
| **Graduate & above** | 3.41±1.63 | 1.20±0.88 | 2.48±1.59 | 1.47±1.49 | 0.92±1.18 |
| **P-Value** | 0.177 | 0.643 | 0.304 | 0.689 | **0.053** |
| ***Marital status*** |  |  |  |  |  |
| **Single** | 3.28±1.09 | 1.27±0.78 | 2.51±1.49 | 1.28±1.55 | 0.88±1.16 |
| **Married** | 3.39±1.44 | 1.16±0.69 | 2.45±1.78 | 1.50±2.33 | 0.74±0.86 |
| **P-Value** | **0.556** | **0.289** | **0.827** | **0.474** | **0.407** |
| ***Socioeconomic status*** |  |  |  |  |  |
| **Low** | 3.43±1.31 | 1.24±0.69 | 2.46±1.82 | 1.55±2.63 | 0.59±0.64 |
| **Middle** | 3.42±1.20 | 1.07±0.31 | 2.39±1.59 | 1.41±2.28 | 0.81±0.81 |
| **High** | 3.30±1.69 | 1.24±0.95 | 2.60±1.87 | 1.49±1.78 | 0.88±1.16 |
| **P-Value** | **0.68** | **0.088** | **0.603** | **0.864** | **0.028** |

**Table 3.** Daily mean intake (servings) of discretionary food items:

| Demographic features | Daily Servings |  | P-Value |
| --- | --- | --- | --- |
|  | Mean | SD |  |
| Overall | 1.05 | 1.13 |  |
| Gender |  |  |  |
| Male | 0.77 | 0.96 | <0.001* |
| Female | 1.28 | 1.22 |  |
| Age Groups |  |  |  |
| 18-30 | 1.2 | 1.12 | 0.27 |
| 30-40 | 1.13 | 1.29 |  |
| 40-50 | 0.93 | 0.98 |  |
| >50 | 0.93 | 1.01 |  |
| Marital Status |  |  |  |
| Single | 1.16 | 0.99 | 0.342 |
| Married | 1.03 | 1.15 |  |
| Education Level |  |  |  |
| Illiterate – primary level | 0.93 | 1.1 | 0.003* |
| Higher secondary | 0.92 | 1.19 |  |
| Graduate and above | 1.31 | 1.04 |  |
| Socio-economic status |  |  |  |
| Low | 0.93 | 1.14 | 0.001* |
| Middle | 0.87 | 1.03 |  |
| High | 1.32 | 1 |  |
| Employment |  |  |  |
| Not Working | 1.18 | 1.12 | 0.049* |
| working | 0.96 | 1.13 |  |

## Discussion

The current study showedthat the dietary intake of the population in major cities of Pakistan do not match with PDGN. Pakistan Dietary guidelines for better nutrition has been developed by the Food and Agriculture Organization of the United Nations and Planning Commission of Pakistan which are almost similar to the dietary guidelines of Eastern Mediterranean Region Office (EMRO) (9) and United States(10) except for cereals (carbohydrates) and fruits. In PDGs, the serving of cereals and fruits are 4-5 and 2-3 while it is 2 and 4 serving in US and EMRO guidelines respectively.

The low adherence to national dietary guidelines is a universal phenomenon (11-15). Similarly, the high intake of carbohydrates is also consistent with the findings from other studies of Pakistan and neighbouring countries (16-18). High intake of carbohydrate is customary in Asia where people eat either wheat or rice daily. The increased intake of carbohydrates has been linked with susceptibility to diabetes mellitus(17, 19) development of metabolic syndrome abnormalities including obesity, dyslipidemia, glucose intolerance and hypertension (20). Therefore, low adherence to the dietary guidelines and high intake of carbohydrate is a matter of concern for the Asian population.

The adherence score of the female participants was high as compared to their counterpart. Similarly, high education level, increasing age and high socioeconomic status were also associated with better adherence. On the other hand, intake of discretionary food items was also significantly high in females, graduate, high socio-economic group. The intake of discretionary foods have been associated with obesity (21), increased risk of depression (22), high cancer and cardiovascular specific mortality (23), increasing risk of asthma (24) and functional gastrointestinal disorders (25).

There are different reasons for low adherence to PDGN. First is the low awareness of dietary guidelines to the public and people are unaware of the importance of a balanced diet. Secondly, it is customary to take more carbohydrates in the diet in our society lacking in vegetables and fruits which lead to a carbohydrate-rich diet. Thirdly, due to urbanization and heavy advertisement, there is a cultural shift to more intakes of discretionary food items instead of fresh fruits/vegetables leading to unhealthy dietary intake behavior.

Pakistan is a developing country facing a double burden of malnutrition and overweight/obesity while a recent survey showed that 40.2% of children are stunted (26). The sustainable development goal 2 is related to nutrition and a balanced diet and to achieve sustainable development goals (SDGs) 2030, there is a need to give due importance to nutrition.There is dire need to revamp the overall nutritional plan and guidelines inPakistanto achieve this goal. A multidisciplinary approach is required involving all key stakeholders including health, education, and media to the public about the importance of a balanced diet. Besides this, proper dissemination of PDGN among public might help in achieving the desired outcome. The current government is sensitized about the importance of nutrition and its impact on health and overall economy of the country and has initiated different programs(27). These include community and health and nutrition initiative, multi-sectoral Nutrition Coordinating Body, National Centre for Human Nutrition, Kitchen Gardening initiative, specialized nutritious food and to address spurious & adulterated milkhowever there is a big challenge in the continuity of these programs.Due to small sample size, the findings of this study cannot be generalized however these provide a strong evidence about the overall adherence pattern in our society.

## Conclusion

The adherence to the national dietary recommendation is poor and need to be addressed by bringing behavior change with the help of information education and communication. Awareness must be created for the selection of healthy dietary food items for themselves and family that will ultimately lead to better health and wellbeing of the society.

## Data Availability

The data will be available.

## Acknowledgments

We acknowledge the efforts of Dr. Huma Qureshi in final revising of the proposal and guidance. The efforts of Liaquat Ali, Amna Bibi, Ms. Bushra Mumtaz, Ms. Najma Pathan, Mr. Masood, Mr. Muhammad Aslam, Amir Zaib Tanoli, Ms. Sana Rehman, Masooma Bukhari, Kaleemullah are acknowledge for assistance in data collection and enumeration. We are also thankful to Mr. Kashif, Muhammad Saqib, Hafiz Saqib, Muhammad Zulfiqar, Waryal Ali Daheri, Khalid Mehmood, Gohar Rehman, Muhamamd Ijaz, Saeed Ahmed Shahid, Sajid Kazmi for data entry.

## Notes

**Financial Support:** This work was supported by WHO Eastern Mediterranean Region Office (RPPH 16-73)

**Conflict of Interest:** The authors declare none conflict of interest.

### Competing Interest Statement

The authors have declared no competing interest.

### Funding Statement

The funding was provided by the WHO.

### Author Declarations

The study was approved by the National Bioethics Committee of Pakistan.

## References

1. Kant AK. Dietary patterns and health outcomes. Journal of the American Dietetic Association. 2004;104(4):615–35.

2. Boylan S, Lallukka T, Lahelma E, Pikhart H, Malyutina S, Pajak A, et al. Socioeconomic circumstances and food habits in Eastern, Central and Western European populations. Public health nutrition. 2011;14(4):678–87.

3. Iqbal R, Tahir S, Ghulamhussain N. The need for dietary guidelines in Pakistan. JPMA The Journal of the Pakistan Medical Association. 2017;67(8):1258–61.

4. 2015–2020 Dietary Guidelines for Americans 2019 [cited 2019 20 august]. Available from: https://health.gov/dietaryguidelines/2015/.

5. Pakistan. World Health Organization. Country Cooperation Strategy 2018 [cited 2019 20 August]. Available from: https://apps.who.int/iris/bitstream/handle/10665/136607/ccsbrief_pak_en.pdf;jsessionid=8E3D74CCA84C373B3856CD5986F42AB1?sequence=1.

6. Rafique I, Saqib MAN, Munir MA, Qureshi H, Rizwanullah, Khan SA, et al. Prevalence of risk factors for noncommunicable diseases in adults: key findings from the Pakistan STEPS survey. Eastern Mediterranean health journal = La revue de sante de la Mediterranee orientale = al-Majallah al-sihhiyah li-sharq al-mutawassit. 2018;24(1):33–41.

7. McLaren L, Sumar N, Barberio AM, Trieu K, Lorenzetti DL, Tarasuk V, et al. Population-level interventions in government jurisdictions for dietary sodium reduction. The Cochrane database of systematic reviews. 2016;9:CD010166.

8. Pakistan Dietary guidelines for better nutrition: Food and Agriculture Organization of the United Nations and Ministry of Planning Development and Reform, Government of Pakistan.

9. Promoting a healthy diet for the WHO Eastern Mediterranean Region: user-friendly guide World Health Organization; 2012.

10. Dietary guidelines 2015-2020.. Available from: https://health.gov/dietaryguidelines/2015/guidelines/chapter-1/about/.

11. Vandevijvere S, De Vriese S, Huybrechts I, Moreau M, Temme E, De Henauw S, et al. The gap between food-based dietary guidelines and usual food consumption in Belgium, 2004. Public health nutrition. 2009;12(3):423–31.

12. de Abreu D, Guessous I, Vaucher J, Preisig M, Waeber G, Vollenweider P, et al. Low compliance with dietary recommendations for food intake among adults. Clinical nutrition. 2013;32(5):783–8.

13. Serra-Majem L, Ribas-Barba L, Salvador G, Serra J, Castell C, Cabezas C, et al. Compliance with dietary guidelines in the Catalan population: basis for a nutrition policy at the regional level (the PAAS strategy). Public health nutrition. 2007;10(11A):1406–14.

14. Al Thani M, Al Thani AA, Al-Chetachi W, Al Malki B, Khalifa SAH, Bakri AH, et al. Adherence to the Qatar dietary guidelines: a cross-sectional study of the gaps, determinants and association with cardiometabolic risk amongst adults. BMC public health. 2018;18(1):503.

15. Zaghloul S, Waslien C, Al Somaie M, Prakash P. Low adherence of Kuwaiti adults to fruit and vegetable dietary guidelines. Eastern Mediterranean health journal = La revue de sante de la Mediterranee orientale = al-Majallah al-sihhiyah li-sharq al-mutawassit. 2012;18(5):461–7.

16. Aziz S, Hosain K. Carbohydrate (CHO), protein and fat intake of healthy Pakistani school children in a 24 hour period. JPMA The Journal of the Pakistan Medical Association. 2014;64(11):1255–9.

17. Mohan V, Unnikrishnan R, Shobana S, Malavika M, Anjana RM, Sudha V. Are excess carbohydrates the main link to diabetes & its complications in Asians? The Indian journal of medical research. 2018;148(5):531–8.

18. Akhter N, Sondhya FY. Nutritional status of adolescents in Bangladesh: Comparison of severe thinness status of a low-income family’s adolescents between urban and rural Bangladesh. Journal of education and health promotion. 2013;2:27.

19. Gross LS, Li L, Ford ES, Liu S. Increased consumption of refined carbohydrates and the epidemic of type 2 diabetes in the United States: an ecologic assessment. The American journal of clinical nutrition. 2004;79(5):774–9.

20. Park S, Ahn J, Kim NS, Lee BK. High carbohydrate diets are positively associated with the risk of metabolic syndrome irrespective to fatty acid composition in women: the KNHANES 2007-2014. International journal of food sciences and nutrition. 2017;68(4):479–87.

21. Joseph N, Nelliyanil M, Rai S, Y PR, Kotian SM, Ghosh T, et al. Fast Food Consumption Pattern and Its Association with Overweight Among High School Boys in Mangalore City of Southern India. Journal of clinical and diagnostic research : JCDR. 2015;9(5):LC13–7.

22. Sanchez-Villegas A, Toledo E, de Irala J, Ruiz-Canela M, Pla-Vidal J, Martinez-Gonzalez MA. Fast-food and commercial baked goods consumption and the risk of depression. Public health nutrition. 2012;15(3):424–32.

23. Barrington WE, White E. Mortality outcomes associated with intake of fast-food items and sugar-sweetened drinks among older adults in the Vitamins and Lifestyle (VITAL) study. Public health nutrition. 2016;19(18):3319–26.

24. Wang CS, Wang J, Zhang X, Zhang L, Zhang HP, Wang L, et al. Is the consumption of fast foods associated with asthma or other allergic diseases? Respirology. 2018;23(10):901–13.

25. Shau JP, Chen PH, Chan CF, Hsu YC, Wu TC, James FE, et al. Fast foods--are they a risk factor for functional gastrointestinal disorders? Asia Pacific journal of clinical nutrition. 2016;25(2):393–401.

26. National Nutrition Survey 2018. Key Findings Report 2018.

27. EHSAAS PROGRAMME PRIME MINISTER’S POLICY STATEMENT. Goverenment of Pakistan 2019 [cited 2019 21 august]. Available from: http://www.pakistan.gov.pk/ehsaas-program.html.

